# Development and validation of the Emotion Regulation Questionnaire – Positive/Negative (ERQ-PN): Does the target of emotion regulation matter?

**DOI:** 10.1101/2024.06.28.24309661

**Authors:** Robinson De Jesús-Romero, José R. Chimelis-Santiago, Lauren A. Rutter, Lorenzo Lorenzo-Luaces

## Abstract

The Emotion Regulation Questionnaire (ERQ) assesses emotion regulation strategies, particularly expressive suppression and cognitive reappraisal. However, the ERQ does not discern between regulating positive vs. negative emotions. Recent research suggests that suppression and reappraisal can impact mental health differently when targeting positive vs negative emotions. We developed and validated the Emotion Regulation Questionnaire - Positive/Negative (ERQ-PN), designed to measure positive and negative forms of suppression and reappraisal strategies. We recruited 963 participants (female = 478) through Prolific.com and administered the ERQ-PN. Participants had an average age of 45 years and were predominantly White (74%) and heterosexual (84%). Structural validity was assessed through confirmatory factor analyses. Model fit was estimated using the comparative fit index and the root-mean-square error of approximation. We also used the Bayesian information criterion to compare the fit of different models. Overall, participants used reappraisal more often to decrease negative emotions (vs. increasing positive) and leaned toward using suppression more for negative (vs. positive) emotions. These analyses revealed that the four-factor model (Model 2) delineating four latent variables (positive reappraisal, negative reappraisal, positive suppression, and negative suppression) had a good fit (RMSEA = 0.07, CFI = 0. 97, TLI = 0.96, *χ*^2^(98) = 531.28, *p* < 0.001). An incremental validity assessment revealed that positive and negative reappraisal correlated similarly with related mental health constructs. By contrast, suppression of negative vs. positive emotions was differentially associated to the validators tested. The ERQ-PN represents a valid measure of emotion regulation that accounts for both positive and negative emotions.

## 1. Introduction

Emotions are essential to human life, influencing our overall well-being. Emotion regulation refers to the ability to manage or modulate the emotions we experience and how we express them [1]. One widely used measure of emotion regulation is the Emotion Regulation Questionnaire (ERQ) [2]. The ERQ assesses two common emotion regulation strategies: reappraisal and suppression. Reappraisal refers to altering how we evaluate a situation to change its emotional impact, while suppression refers to inhibiting the outward expression of an emotion [1]. In the process model of emotion regulation proposed by Gross [1], these strategies encapsulate two different aspects of the process, namely cognitive change vs. modulation of behavioral and physiological response. However, research has shown that reappraisal and suppression can be used to target positive or negative emotions separately, with distinctive effects on mental health [3,4]. The current edition of the ERQ was not developed to capture the difference between positive and negative emotions, therefore it does not distinguish between these two emotion regulation targets. This paper proposes and evaluates a revision of the ERQ to include questions for measuring both positive and negative reappraisal and suppression strategies, by only adding a handful of questions and addressing a limitation of the ERQ, the different number of items per subscale.

### 1.1. Negative and Positive Cognitive Reappraisal

Reappraisal to up-regulate positive emotions (e.g., “I should feel proud of myself; I reached the best decision possible.”) vs. down-regulate negative ones (e.g., “I should not feel bad about this; I reached the best decision possible”) has been used interchangeably in previous literature. Previous studies have found that reappraising to down-regulate negative emotions can lead to positive psychological outcomes, on average. For example, in a study using ecological momentary assessment, Southward et al. [3] found that participants who used reappraisal of specific negative emotions, such as anger and sadness, could improve their mood. An alternate approach to improving mood involves up-regulating positive emotions instead of down-regulating negative emotions. For example, past research has explored the distinction between these two emotion regulation targets using daily journals [5]. Researchers observed that reappraising to up-regulate emotions was linked to higher positive affect, self-esteem, and psychological adjustment. However, they found no strong association between reappraisal to down-regulate negative emotions and self-esteem or psychological adjustment, suggesting that there may be a distinction between downregulating negative emotions and up-regulating positive emotions.

### 1.2. Negative and Positive Expressive Suppression

Suppression has been studied mainly in the context of suppressing negative emotions [6]. Multiple studies have associated suppression of negative emotions with increased negative affect and depressive symptoms [5,7,8]. For instance, in an experimental study, when participants were asked to suppress their negative emotions to an emotional clip or to allow their emotional response, those that suppressed reported higher distress and heart rate variability [9]. This suggests that suppression may be counterproductive when trying to manage negative emotions. However, depending on the context, suppressing negative emotions can be beneficial. For example, a study exploring suppression of anger in romantic relationships found that suppression of anger when participants were experiencing high (but not low) heart rate variability was associated with better relationship outcomes [10].

Similarly, suppressing positive emotions can have beneficial or harmful effects depending on the context. For example, suppression of positive emotions could lead to higher stress levels, as there is evidence that positive emotions have a stress-buffering effect [11], while expressing positive emotions has been shown to be essential for developing and maintaining social relationships [12]. Thus, suppressing positive emotions could lead to adverse social outcomes such as a higher sense of inauthenticity, which was associated in previous studies with lower social relationship satisfaction and social support [13]. Furthermore, not suppressing positive emotion can also be harmful when it leads to positive-emotion persistence which is believed to be an underlying mechanism in mania [14]. Thus, being able to study positive suppression (separate from negative suppression) could allow us to explore different associations with mental health-related variables.

### 1.3. Our Study

In the current study, we revised the ERQ to assess positive and negative forms of reappraisal and suppression strategies. We define positive reappraisal as the reinterpretation of the meaning of a situation with the goal of up-regulating positive emotions. Negative reappraisal was defined as the reinterpretation of an event with the goal of down-regulating negative emotions. Similarly, we define negative suppression as inhibiting the expression of negative emotions, while positive suppression involves inhibiting the expression of positive emotions. We also explore the associations between these four emotion regulation strategies and different validators related to mental health. Study data and code can be accessed on the OSF website (https://osf.io/ezngp).

## 2. Materials and Methods

### 2.1. Participants

A total of 963 participants were included in this study from a nationally representative sample recruited through Prolific.com, an online participant recruitment platform that has consistently been found to produce better data quality than alternatives like Amazon Mechanical Turk [15]. Recruitment spanned from January 20, 2023, to January 28, 2023. We aimed for a sample of 1000, Prolific’s recommendation for a nationally representative sample based on stratification across three demographics: age, sex and race-ethnicity based on the US Census Bereau [16,17]. We ultimately recruited 963 individuals for the study. This sample size would allow us to detect effect of *r* = 0.09, with power of 80% at *p* value < 0.05.

### 2.2. Measures

#### 2.2.1. Cognitive Reappraisal and Expressive Suppression

Participants’ habitual use of reappraisal and suppression was assessed using the original Emotion Regulation Questionnaire (ERQ). The ERQ has two subscales measuring reappraisal and suppression. The reappraisal 6-item subscale assesses participants’ habitual use of reappraisal by asking participants how much they agree with specific statements on a 7-point scale (1 = strongly disagree, 7 = strongly agree). Mean scores range from 1 to 7, with higher scores representing higher habitual use of suppression. The suppression 4-item subscale assesses participants’ habitual use of suppression by asking participants how much they agree with specific statements on a 7-point scale (1 = strongly disagree, 7 = strongly agree). Mean scores range from 1 to 7, with higher scores representing higher habitual use of suppression. The ERQ demonstrated to have possess high acceptable consistency (𝜔 = 0.75)

#### 2.2.2. Positive and Negative Cognitive Reappraisal and Expressive Suppression

Positive and negative emotion regulation was assessed using the using the ERQ-PN. The ERQ-PN is 16 items measure of four proposed variation of emotion regulation strategies: positive reappraisal (e.g., “I control my positive emotions by changing the way I think about the situation I’m in.”), negative reappraisal (e.g., “I control my negative emotions by changing the way I think about the situation I’m in.”), positive suppression (e.g., “I keep my positive emotions to myself”), and negative suppression (e.g., “I keep my negative emotions to myself.”) Additionally, the instructions and some items were adapted to include examples of negative (i.e., sadness, anger, anxiety, or fear), and positive emotion (i.e., joy, happiness, or positive surprises).

We included four items for each of the four subscales. Thus, the ERQ-PN addresses a limitation of the original ERQ, that the reappraisal scale was assessed with more items that the suppression scale. The ERQ-PN items were adapted from the ERQ by including a specific type of emotional target (i.e., positive vs negative emotion). However, we retained much of the same language in the items so that the positive and negative versions were textually “mirror” versions of each other. When the original ERQ items were about positive emotions (e.g., “When I want to feel more positive emotion (such as joy or amusement), I change what I’m thinking about”), we reused the item and added one that focused on negative emotions (i.e., “When I want to feel less negative emotions (e.g., sadness, anger, anxiety, or fear), I change what I’m thinking about.” and vice versa. When items did not specify type of emotion (e.g., “I keep my emotions to myself.”), we modify it to include positive or negative emotions (e.g., “I keep my positive emotions to myself.”) We also matched the number of items for all the subscales, so that each subscale would have four items. This meant that we excluded one of the ERQ reappraisal items (e.g., “I control my emotions by changing the way I think about the situation I’m in.”) We found this item would be redundant with other reappraisal items (e.g., “When I want to feel more positive emotion, I change the way I’m thinking about the situation.”, “When I want to feel more positive emotion, I change the way I’m thinking about the situation.”) The items were also organized in the same order as the ERQ; however, we began with all items for positive reappraisal and suppression, followed by all items for negative reappraisal and suppression. Participants rated each item on a seven-point Likert scale, ranging from 1 (strongly disagree) to 7 (strongly agree). Mean scores range from 1 to 7, with higher scores representing higher habitual use of the specific emotion regulation strategy. A copy of the ERQ-PN measure can be found on appendix B. The ERQ-PN demonstrated to have high internal consistency (𝜔 = 0.84)

#### 2.2.3. Difficulties in Emotion Regulation

Participants’ difficulties in emotion regulation were assessed using the brief version of the Difficulties in Emotion Regulation Scale (DERS-18) [18,19]. This 18-item scale measures six aspects of emotion regulation: nonacceptance (i.e., nonacceptance of emotional responses), goals (i.e., difficulty engaging in goal-directed behavior), impulse (i.e., impulse control difficulties), awareness (lack of emotional awareness), strategies (i.e., limited access to emotion regulation strategies), and clarity (i.e., lack of emotional clarity). Participants were asked to indicate how often the items applied to them on a 5-point Likert scale (1 = almost never, 5 = almost always).

Scores range from 3 to 15 for each subscale, with higher scores indicating greater difficulties in emotion regulation. The DERS-18 demonstrated to possess excellent internal consistency (𝜔 = 0.91)

#### 2.2.4. Psychological Distress

Participants’ level of internalizing distress was measured using the Kessler Psychological Distress Scale (K6) [20]. The K6 is a 6-item scale that assesses internalizing distress by asking participants to rate on a 4-point scale how often they have experienced negative affect and related symptoms over the past month (0 = none of the time, 4 = all of the time). Scores range from 0 to 24, with higher scores indicating higher psychological distress. Scores of 6 on the K6 may indicate mild psychological distress, while scores of 13 may indicate more severe psychological distress. The K6 has been demonstrated to have criterion validity [21] and in our study demonstrated to possess high internal consistency (𝜔 = 0.87)

#### 2.2.5. Well-being

Participants’ subjective well-being was assessed using the World Health Organization Well-being Index (WHO-5) [22]. This 5-item scale asks participants to rate their experiences based on their feelings over the last two weeks on a 6-point Likert scale (0 = at no time, 5 = all of the time). Scores range from 0 to 25 and are multiplied by four to produce scores from 0-100, with higher scores representing higher well-being. The WHO-5 demonstrated to have excellent internal consistency (𝜔 = 0.92).

#### 2.2.6. Positive and Negative Affect

Participants’ positive and negative affect were assessed using the Positive and Negative Affect Schedule (PANAS) [23]. This 20-item scale consists of two subscales: positive affect and negative affect, each containing 10 items. Participants were asked to rate the extent to which they have felt each emotion on a 5-point Likert scale (1 = very slightly or not at all, 5 = extremely).

Scores range from 10 to 50, with higher scores indicating higher positive or negative affect levels. The PANAS demonstrated to have acceptable internal consistency (𝜔 = 0.72).

#### 2.2.7. Personality

We assessed the Big-Five personality traits (i.e., emotional stability, extraversion, agreeableness, conscientiousness, and openness) using the Ten-Item Personality Inventory (TIPI) [24]. The TIPI is a 10-item scale assessing personality traits with 5 bipolar factors representing extraversion, agreeableness, conscientiousness, emotional stability, and openness to experience. The measure contains 2 descriptors for each pole of all 5 personality dimensions. Each item is rated using a 7-point scale (1 = disagree strongly, 7 = agree strongly). After reverse recoding, the mean for each of the 5 personality dimensions was used as subscales. Scores range from 1 to 7, with higher scores indicating higher endorsement of the personality trait. The TIPI demonstrated to have acceptable internal consistency (𝜔 = 0.76).

### 2.3. Procedure

Participants who wished to partake in the study were presented with a Qualtrics survey. On the first page of the survey, participants were provided with a Study Information Sheet, which detailed the study’s purpose, procedures, potential risks and benefits, confidentiality, and payment (2.40 USD). Consent was obtained by participants clicking on a button to indicate their agreement and desire to proceed. Following consent, participants were presented with demographic questions. Subsequently, they were given the original ERQ and the ERQ-PN. To control for familiarity with the questionnaires, the order of these measures was randomized. An explanation was included between the first and second ERQ, informing participants that the next questionnaire would be very similar and requesting participants to complete the questionnaire the same way that they would answer any other. Upon completing these two questionnaires, participants proceeded to complete the remaining measures.

### 2.4. Analytic Plan

All analyses were conducted using the R programming language [25]. We first conducted descriptive statistics for demographic and psychological factors. We then conducted descriptive statistics for the ERQ-PN including mean, standard deviation, median, and interquartile range.

### 2.5. Model Fit

To evaluate the consistency of our theoretical model with the observed data, we employed confirmatory factor analysis (CFA), a statistical method used to test the hypothesized relationship between observed variables (i.e., ERQ-PN items) and the predicted underlying latent constructs (positive vs negative reappraisal and suppression). We first tested a two-factor model (Model 1), where we attempted to replicate the traditional two-factor ERQ structure in which the items were loaded onto one of two latent variables representing different emotion regulation strategies: reappraisal or suppression. Next, we tested a four-factor model (Model 2) with four correlated latent variables: positive reappraisal, negative reappraisal, positive suppression, and negative suppression. we examined a second-order model (Model 3), adding two higher-order latent variables representing overall reappraisal (i.e., positive and negative reappraisal) and suppression (i.e., positive and negative suppression).

### 2.6. Model Comparison

To assess the degree to which the models fit the data, we relied on the comparative fit index (CFI) [26] and the root-mean-square error of residual approximation (RMSEA) [27].

Although guidelines for good fit may vary, generally a CFI value above .95 and RMSEA values less than 0.06 represent a good fit to the data [28]. We also used the Bayesian information criterion (BIC) to identify which of the good-fitting models was most parsimonious. When comparing models, a lower BIC value implies a better fit of the data in terms of the odds of the model, with the lowest BIC value being superior to other models [29]. Additionally, we conducted a nested chi-squared test to identify whether the more complex model significantly improved fit compared to the simpler one [30].

### 2.7. Convergent Validity and Concurrent Validity

Finally, we explored the correlation between the ERQ and ERQ-PN subscales, and dimensions of positive and negative mental health to establish convergent validity. Specifically, we correlated the two ERQ subscales (i.e., suppression and reappraisal), and the ERQ-PN four sub-scales (i.e., positive reappraisal, negative reappraisal, positive suppression, and negative suppression) with 12 validators reflecting positive and negative dimensions of mental health.

Additionally, we used Zou’s test for dependent overlapping correlations [31] to evaluate the difference between positive and negative versions of reappraisal and suppression. We used this because it allowed us to test whether these two pairs of correlations significantly differ from each other. This test takes into account the fact that the correlations are not independent of each other (e.g., (1) negative reappraisal and well-being, (2) positive reappraisal and well-being). The Zou’s test also allowed us to examine the size of the differences and statistical significance at *p* < .05.

## 3. Results

### 3.1. Demographics

The study included a sample of U.S. adults (N = 963) meant to be nationally representative in terms of age, sex assigned at birth, and race-ethnicity. Sample demographics are found in Table 1.

**Table 1.** Demographic and psychological characteristics of a nationally representative sample of participants who participated in an online study to validate an adapted version of the Emotion

| **Variable** | **N (%)** |
| --- | --- |
| **Gender** |  |
| Woman | 478 (49.69%) |
| Man | 467 (48.54%) |
| Non-binary or genderqueer | 14 (1.46%) |
| Other | 3 (0.31%) |
| Missing | 1 |
| **Race/Ethnicity** |  |
| Non-Hispanic White | 709 (74.01%) |
| Non-Hispanic Black | 115 (12.00%) |
| Hispanic | 59 (6.16%) |
| Asian | 55 (5.74%) |
| Multiracial | 14 (1.46%) |
| American Indian/Native American or Alaska Native | 3 (0.31%) |
| Other | 2 (0.21%) |
| Native Hawaiian or Other Pacific Islander | 1 (0.10%) |
| Missing | 5 |
| **Sexual orientation** |  |
| Heterosexual | 810 (84.20%) |
| Bisexual | 87 (9.04%) |
| Homosexual/Gay/Lesbian | 36 (3.74%) |
| Asexual | 17 (1.77%) |
| Other | 12 (1.25%) |
| Missing | 1 |
| **Education** |  |
| Some high school or less | 6 (0.63%) |
| High school diploma or GED | 133 (13.87%) |
| Some college, but no degree | 176 (18.35%) |
| Associate or technical degree | 109 (11.37%) |
| Bachelor's degree | 373 (38.89%) |
| Graduate or professional degree (MA, MS, MBA, PhD, JD, MD, DDS, etc.) | 162 (16.89%) |
| Missing | 4 |
| **Yearly income** |  |
| Under \$15,000 | 72 (7.51%) |
| \$15,000 - \$24,999 | 92 (9.59%) |
| \$25,000 - \$34,999 | 96 (10.01%) |
| \$35,000 - \$49,999 | 134 (13.97%) |
| \$50,000 - \$74,999 | 212 (22.11%) |
| \$75,000 - \$99,999 | 140 (14.60%) |
| \$100,000 - \$149,999 | 122 (12.72%) |
| \$150,000 - \$199,999 | 55 (5.74%) |
| \$200,000+ | 36 (3.75%) |
| Missing | 4 |
| **Age (in years)** | 45.46 (15.64) |
| Missing | 5 |
| **Reappraisal of positive emotions (ERQ-PN; 1-7)** | 4.79 (1.26) |
| **Suppression of positive emotions (ERQ-PN; 1-7)** | 2.43 (1.31) |
| **Reappraisal of negative emotions (ERQ-PN; 1-7)** | 5.05 (1.24) |
| **Suppression of negative emotions (ERQ-PN; 1-7)** | 4.08 (1.58) |
| **Cognitive reappraisal (ERQ; 1-7)** | 5.03 (1.16) |
| Missing | 7 |
| **Expressive suppression (ERQ; 1-7)** | 3.47 (1.35) |
| Missing | 9 |
| **Extraversion (TIPI; 1-7)** | 3.54 (1.76) |
| Missing | 6 |
| **Agreeableness (TIPI; 1-7)** | 5.47 (1.24) |
| Missing | 2 |
| **Conscientiousness (TIPI; 1-7)** | 5.52 (1.34) |
| Missing | 2 |
| **Emotional stability (TIPI; 1-7)** | 4.76 (1.65) |
| Missing | 1 |
| **Openness (TIPI; 1-7)** | 5.17 (1.30) |
| Missing | 2 |
| **Awareness (DERS-18; 3-15)** | 6.50 (2.54) |
| Missing |  |
| **Clarity (DERS-18; 3-15)** | 5.07 (2.28) |
| Missing | 6 |
| **Goals (DERS-18; 3-15)** | 8.83 (3.52) |
| Missing | 10 |
| **Impulse (DERS-18; 3-15)** | 4.93 (2.60) |
| Missing | 9 |
| **Nonacceptance (DERS-18; 3-15)** | 6.58 (3.43) |
| Missing | 7 |
| **Strategies (DERS-18; 3-15)** | 6.05 (3.12) |
| Missing | 8 |
| **Difficulty in emotion regulation (DERS; 18-90)** | 37.92 (12.50) |
| Missing | 36 |
| **Positive affect (PANAS; 10-50)** | 26.92 (7.70) |
| Missing | 19 |
| **Negative affect (PANAS; 10-50)** | 17.25 (5.60) |
| Missing | 20 |
| **Internalizing distress (K6;0-24)** | 5.30 (5.11) |
| Missing | 9 |
| **Well-being (WHO-5; 0-100)** | 54.19 (23.47) |
| Missing | 7 |

*Note.* N = total sample size, M = mean score, S.D. = standard, % = percentage. ERQ = Emotion Regulation Questionnaire, ERQ-PN = revised ERQ, TIPI = Ten Item Personality Inventory, DERS-18 = Brief version of the Difficulties in Emotion Regulation Scale, PANAS = Positive and Negative Affect Scale, K6 = Kessler Psychological Distress Scale, WHO-5 = World Health Organization Well-Being Index. Positive values = positive correlation, negative values = negative correlation. Ranges are shown within parentheses for the continuous variables.

### 3.2. ERQ-PN

Descriptive statistics for the ERQ-PN items are presented in Table 2. In general, participants reported greater use of reappraisal to feel less negative (e.g., sad, angry, anxious, or fearful) than to feel more positive (e.g., joy, happiness, or surprise; *t*(962) = -8.53, *p* < .001, mean difference = -0.26, 95% *CI* [-0.32, -0.20], *d* = -0.27). Participants appeared to use suppression much more with negative than with positive emotions (*t*(962) = -32.94, *p* < .001, mean difference = -1.65, 95% *CI* [-1.75, -1.55], *d* = -1.06).

**Table 2.** Descriptive statistics of the Revised Emotion Regulation Questionnaire’s latent factors and specific items in a sample of 963 adults recruited via Prolific (N = 963)

| **Variables** | **Mean**<br>**(Range: 1-7)** | **SD** | **Median** | **IQR**<br>**25%** | **IQR**<br>**75%** |
| --- | --- | --- | --- | --- | --- |
| **Subscales** |  |  |  |  |  |
| Reappraisal of positive emotions | 4.79 | 1.26 | 5.00 | 4.00 | 5.75 |
| Suppression of positive emotions | 2.43 | 1.31 | 2.25 | 1.25 | 3.25 |
| Reappraisal of negative emotions | 5.05 | 1.24 | 5.00 | 4.50 | 6.00 |
| Suppression of negative emotions | 4.08 | 1.58 | 4.25 | 3.00 | 5.25 |
| **Items** |  |  |  |  |  |
| 1: "When I want to feel less negative emotions (e.g., sadness, anger, anxiety, or fear), I change what I'm thinking about." | 5.13 | 1.40 | 5.00 | 5.00 | 6.00 |
| 2: "I keep my negative emotions to myself." | 4.33 | 1.71 | 5.00 | 3.00 | 6.00 |
| 3: "When I am feeling negative emotions, I am careful not to express them." | 4.20 | 1.68 | 4.00 | 3.00 | 5.00 |
| 4: "When I'm faced with a stressful situation, I make myself think about it in a way that helps me feel less negative (e.g., less sad, angry, anxious, or fearful)." | 4.98 | 1.41 | 5.00 | 4.00 | 6.00 |
| 5: "I control my negative emotions by not expressing them." | 3.84 | 1.77 | 4.00 | 2.00 | 5.00 |
| 6: "When I want to feel less negative emotion, I change the way I'm thinking about the situation." | 5.07 | 1.37 | 5.00 | 5.00 | 6.00 |
| 7: "I control my negative emotions by changing the way I think about the situation I'm in." | 5.00 | 1.40 | 5.00 | 4.00 | 6.00 |
| 8: "When I am feeling negative emotions, I make sure not to express them." | 3.96 | 1.75 | 4.00 | 3.00 | 5.00 |
| 9: "When I want to feel more positive emotion (e.g., joy, happiness, or surprise), I change what I'm thinking about." | 4.97 | 1.55 | 5.00 | 4.00 | 6.00 |
| 10: "I keep my positive emotions to myself." | 2.68 | 1.50 | 2.00 | 1.00 | 4.00 |
| 11: "When I am feeling positive emotions, I am careful not to express them." | 2.36 | 1.35 | 2.00 | 1.00 | 3.00 |
| 12: "When I'm faced with a stressful situation, I make myself think about it in a way that helps me feel more positive (e.g., joyful, happy, surprised)." | 4.91 | 1.47 | 5.00 | 4.00 | 6.00 |
| 13: "I control my positive emotions by not expressing them." | 2.38 | 1.42 | 2.00 | 1.00 | 3.00 |
| 14: "When I want to feel more positive emotion, I change the way I'm thinking about the situation." | 4.91 | 1.52 | 5.00 | 4.00 | 6.00 |
| 15: "I control my positive emotions by changing the way I think about the situation I'm in." | 4.37 | 1.67 | 5.00 | 3.00 | 6.00 |
| 16: "When I am feeling positive emotions, I make sure not to express them." | 2.29 | 1.39 | 2.00 | 1.00 | 3.00 |

*Note.* *SD* = standard, *IQR* = interquartile range.

### 3.3. Structural Validity

The model with two latent variables representing reappraisal and suppression (Model 0) fit the data poorly (RMSEA = 0.19, CFI = 0.70, TLI = 0.65, *χ*^2^(103) = 3837.29, *p* < 0.001), suggesting that in this sample, variation in the scale is not well-explained by these two factors. The four-factor model (Model 1) representing four correlated latent variables, positive reappraisal, negative reappraisal, positive suppression, and negative suppression (see Fig 1), fit the data well. However, the *χ*^2^ did not meet conventional cut-offs (RMSEA = 0.07, CFI = 0. 97, TLI = 0.96, *χ*^2^(98) = 531.28, *p* < 0.001). Finally, the second-order model (Model 2), which included two higher-order latent variables representing overall reappraisal (i.e., positive and negative reappraisal) and suppression (i.e., positive and negative suppression, see Fig 2) also fit the data well (RMSEA = 0.07, CFI = 0.96, TLI = 0.96, *χ*^2^(101) = 563.29, *p* < 0.001). For this model we fixed to 1 the first order latent to reduce overparameterization. Model 1 exhibited lower AIC and BIC values (AIC = 31440, BIC = 31625) than Model 2 (AIC = 31466, BIC = 31636), indicating that Model 1 is more parsimonious than Model 2. These results suggest that the items we added to the ERQ may capture the intended differences in positive vs. negative emotion regulation, although falling under the umbrella of habitual use of reappraisal and suppression. Factor loadings can be found on appendix C.

**Fig 1.**
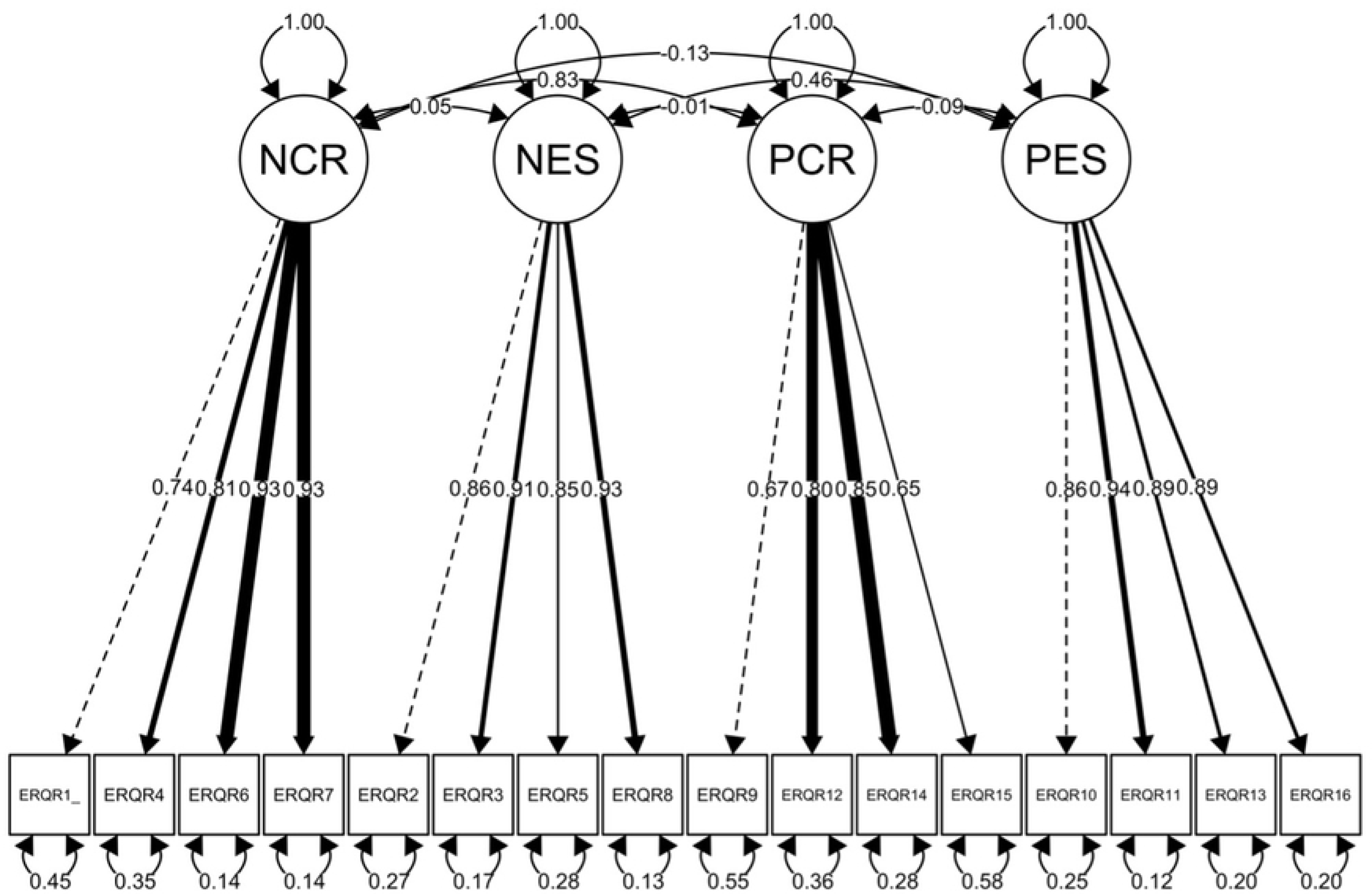
Structural Confirmatory Factor Analysis of Four-Factor Model: Negative Reappraisal, Negative Suppression, Positive Reappraisal, and Positive Suppression in a sample of Online Respondents (N = 963) *Note*. The figure represents a confirmatory factor analysis (CFA) model with four latent variables, NCR = negative cognitive reappraisal, NES = negative expressive suppression, PCR = positive cognitive reappraisal, and PES = positive expressive suppression, derived from the ERQ-PN scale. Observed variables, represented as ERQR1 through ERQR16, are individual items on the ERQ-PN scale. Factor loadings, error variances, and correlations between the factors are also depicted.

**Fig 2.**
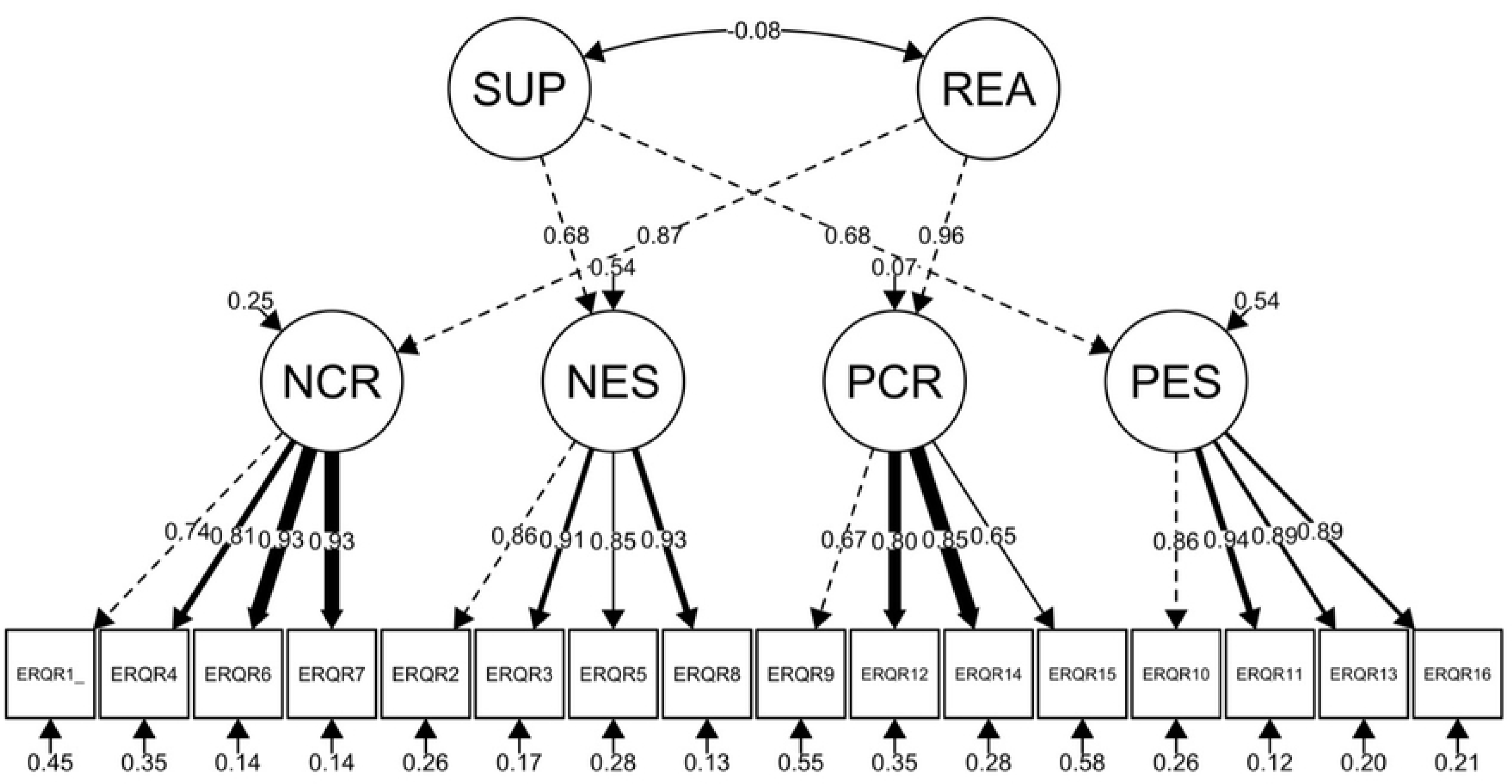
Structural Confirmatory Factor Analysis of Higher Order Positive and Negative Emotion Traits: Positive Reappraisal, Suppression, Negative Reappraisal, and Suppression (N = 963) *Note*. The figure represents a Confirmatory Factor Analysis (CFA) model with six latent variables: SUP = positive and negative suppression, REA = positive and negative reappraisal, NCR = negative cognitive reappraisal, NES = negative expressive suppression, PCR = positive cognitive reappraisal and PES = positive expressive suppression, derived from the ERQ-PN scale. Observed variables, represented as ERQR1 through ERQR16, are individual items on the ERQ-PN scale. Factor loadings, error variances, and correlations between the factors are also depicted.

### 3.4. Convergent Validity and Concurrent Validity

To evaluate the convergent and concurrent validity of the ERQ-PN’s distinction between positive and negative emotion regulation over the traditional ERQ, we correlated the four emotion regulation sub-scales (i.e., positive reappraisal, negative reappraisal, positive suppression, and negative suppression) and the original ERQ subscales with various constructs reflecting positive and negative dimensions of mental health (see Table 3). The table suggests that the ERQ-PN has convergent validity as most of the correlations between negative/positive reappraisal and suppression, follow a similar pattern that the correlations with their ERQ reappraisal and suppression counterparts.

**Table 3.** Correlations Between Emotion Regulation Strategies and Mental Health Outcomes in a Nationally Representative Sample of Participants (N = 963)

|  | Cognitive<br>reappraisal<br>(ERQ) | Reappraisal<br>of positive<br>emotions<br>(ERQ-PN) | Reappraisal<br>of negative<br>emotions<br>(ERQ-PN) | Expressive<br>suppression<br>(ERQ) | Suppression<br>of positive<br>emotions<br>(ERQ-PN) | Suppression<br>of negative<br>emotions<br>(ERQ-PN) |
| --- | --- | --- | --- | --- | --- | --- |
| Extraversion<br>(TIPI) | 0.15*** | 0.14*** | 0.17*** | -0.36*** | -0.28*** | -0.26*** |
| Emotional<br>stability (TIPI) | 0.35*** | 0.35*** | 0.38*** | -0.01 | -0.06 | 0.08* |
| Awareness<br>(DERS-18) | -0.31*** | -0.29*** | -0.29*** | 0.36*** | 0.27*** | 0.28*** |
| Clarity<br>(DERS-18) | -0.28*** | -0.23*** | -0.27*** | 0.26*** | 0.26*** | 0.15*** |
| Goals (DERS-18) | -0.19*** | -0.19*** | -0.23*** | 0.06 | -0.01 | 0.01 |
| Impulse<br>(DERS-18) | -0.26*** | -0.25*** | -0.27*** | 0.03 | 0.11** | -0.04 |
| Nonacceptance<br>(DERS-18) | -0.18*** | -0.15*** | -0.17*** | 0.26*** | 0.20*** | 0.22*** |
| Strategies<br>(DERS) | -0.35*** | -0.34*** | -0.38*** | 0.15*** | 0.17*** | 0.07* |
| Difficulty in<br>emotion<br>regulation<br>(DERS-18) | -0.36*** | -0.34*** | -0.37*** | 0.24*** | 0.22*** | 0.15*** |
| Positive affect<br>(PANAS) | 0.37*** | 0.36*** | 0.38*** | -0.21*** | -0.20*** | -0.09** |
| Negative affect<br>(PANAS) | -0.22*** | -0.2*** | -0.24*** | 0.12** | 0.1** | 0.03 |
| Internalizing<br>distress (K6) | -0.29*** | -0.27*** | -0.30*** | 0.21*** | 0.19*** | 0.12*** |
| Well-being<br>(WHO-5) | 0.32*** | 0.32*** | 0.35*** | -0.18*** | -0.17*** | -0.07* |

*Note.* Table values are Pearson's $r$ between the ERQ, ERQ-PN, TIPI, DERS-18, PANAS, K6, and WHO-5. ERQ = Emotion Regulation Questionnaire, ERQ-PN = ERQ Revised, TIPI = Ten Item Personality Inventory, DERS-18 = Brief version of the Difficulties in Emotion Regulation Scale, PANAS = Positive and Negative Affect Scale, K6 = Kessler Psychological Distress Scale,
WHO-5 = World Health Organization Well-Being Index. \*\*\* = $p < 0.001$ , \*\* = $p < 0.01$ , \* = $p < 0.05$ .

The data also supported some degree of convergent validity in assessing positive vs. negative emotion regulation, at least for suppression. Notably, positive and negative reappraisal showed differing correlations with the 4 of the 12 validators (see Figure 3). The suppression sub-scales exhibited larger differential associations with 6 of the 12 validators (see Figure 4). For example, difficulties with impulse control were positively correlated with positive suppression *(r* = 0.11) and negatively, though weakly, associated with negative suppression (*r* = -0.04), which represented a statistically significant difference (*Z* = 4.53, *p* < 0.001). Notably, positive reappraisal was highly correlated with negative reappraisal (*r* = 0.89; *CI* [0.88, 0.90]). However, the confidence intervals did not reach 1 suggesting that they are different terms.

**Fig 3.**
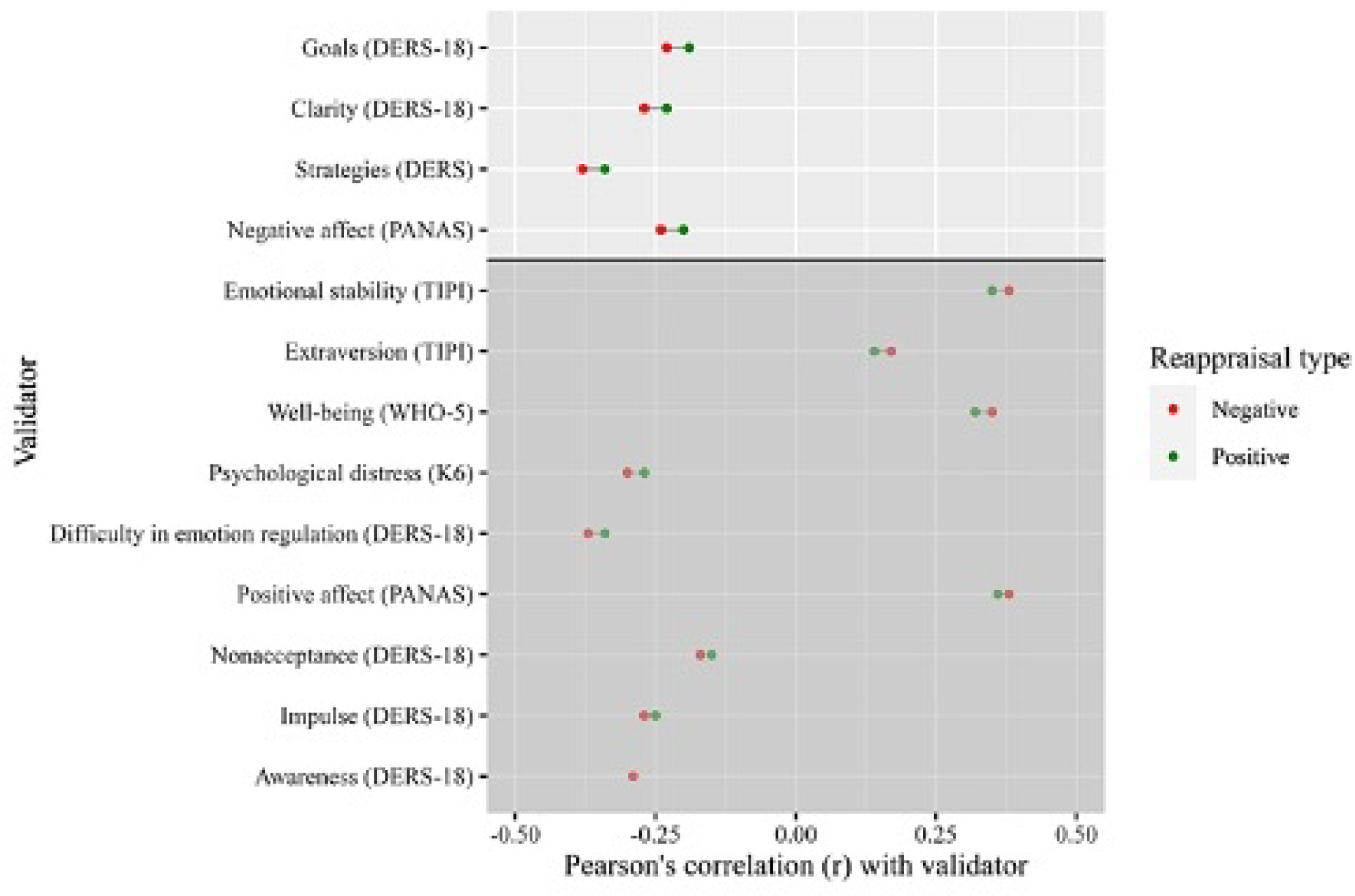
Convergent validity of negative vs positive reappraisal, across different validators in a nationally representative sample of online workers (N = 963) *Note*. Figure values are Pearson’s r between the ERQ-PN and validators. Values displayed above the horizontal line indicate differences in Pearson’s correlations of 0.04 or greater, which are statistically significant at *p* < 0.05 according to Zou’s tests. ERQ = Emotion Regulation Questionnaire, ERQ-PN = Emotion Regulation Questionnaire Positive and Negative, TIPI = Ten Item Personality Inventory, DERS-18 = Brief version of the Difficulties in Emotion Regulation Scale, PANAS = Positive and Negative Affect Scale, K6 = Kessler Psychological Distress Scale, WHO-5 = World Health Organization Well-Being Index.

**Fig 4.**
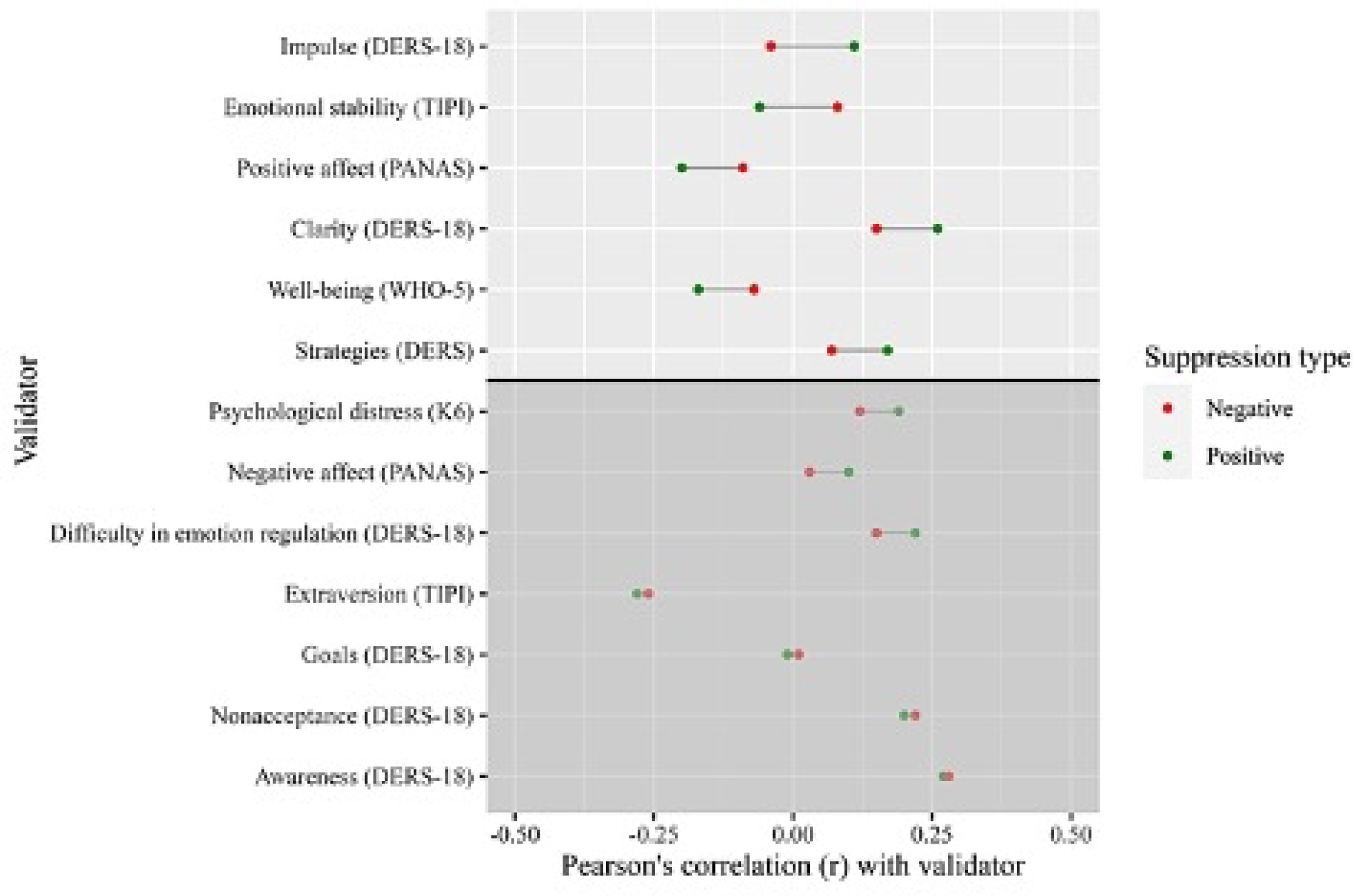
Convergent validity of negative vs positive suppression, across different validators in a nationally representative sample of online workers (N = 963) *Note*. Figure values are Pearson’s r between the ERQ-PN and validators. Values displayed above the horizontal line indicate differences in Pearson’s correlations of 0.10 or greater, which are statistically significant at *p* < 0.05 according to Zou’s test. ERQ = Emotion Regulation Questionnaire, ERQ-PN = Emotion Regulation Questionnaire Positive and Negative, TIPI = Ten Item Personality Inventory, DERS-18= Brief version of the Difficulties in Emotion Regulation Scale, PANAS = Positive and Negative Affect Scale, K6 = Kessler Psychological Distress Scale, WHO-5 = World Health Organization Well-Being Index.

## 4. Discussion

We aimed to develop and provide preliminary validation for the Emotion Regulation Questionnaire - Positive/Negative (ERQ-PN), which encompasses the regulation of both positive and negative emotions. The best-fitting structure contained latent variables representing reappraisal and suppression, with lower-order factors representing the regulation of negative vs. positive emotions. This model extends the original ERQ by incorporating a potentially important distinction between positive and negative emotion targets. Participants used reappraisal and suppression more often to regulate negative emotions vs. positive ones. Interestingly, positive and negative reappraisal demonstrated high inter-correlations and consistent associations with external validators. In contrast, the positive vs negative suppression items exhibited more differential relations when compared to each other. For instance, positive suppression correlated with increased difficulty with impulsive control, an association not observed with negative suppression. Similarly, positive suppression had stronger associations with well-being and positive affect than negative suppression.

### 4.1. Differences Between Reappraisal and Suppression

These findings advance research in emotion regulation by introducing a measurement tool and highlighting the role of emotional valence in the study of emotion regulation and its association with mental health outcomes. In the current study, distinguishing between positive and negative suppression appeared more meaningful than distinguishing positive from negative reappraisal. Moreover, the associations between positive vs negative reappraisal and mental health outcomes did not markedly differ, suggesting a potential overlap between the two reappraisal subscales. The largest distinction between positive and negative reappraisal was found in variables that were more strongly correlated with positive than negative reappraisal (i.e., lack of emotional clarity, difficulty engaging in goal-directed behavior, and limited access to emotion regulation strategies). However, these differences were of small magnitude.

The overlap between positive and negative reappraisal might be partially explained by the inherent differences between attempting to change cognition vs emotional expression, which impacts emotion regulation at different points of the regulatory process [1]. Specifically, reappraisal aims to alter one’s perception of a situation with the goal of changing the intensity of an emotion. In contrast, suppression primarily focuses on changing the outward expression of emotions, not necessarily the emotion itself. Such modulation may be particularly relevant in specific social contexts where expressing specific emotions is expected (e.g., smiling after receiving a compliment), even if they do not align with the individual’s current emotions. Conversely, reappraisal seeks to adjust the intensity or nature of the emotional experience itself. This distinction, especially when considering the valence of emotions, warrants further exploration.

### 4.2. Differences Between Positive and Negative Suppression

Taken together, our results suggest that suppression of positive emotions could be more closely associated with negative aspects of mental health than negative suppression. Specifically, higher suppression of the expression of negative emotions was positively associated with higher emotional stability. In contrast, suppression of positive emotions was positively associated with higher impulse control difficulties when feeling upset, lower well-being and positive affect, and difficulties with emotion regulation including having limited access to emotion regulation strategies.

Previous experimental research suggests that suppression does not impact positive affect directly when watching an emotion-eliciting film [32]. However, other research shows that suppression of positive emotions can disrupt emotional processing in social interactions [33], potentially leading to reduced well-being and positive affect. Suppressing positive emotions in social situations where an expression of positive affect is expected can lead to diminished social bonds, as the expression of positive emotions plays an essential role in rapport building [34,35].

We also found that individuals with high impulse control difficulties tend to suppress positive emotions more often. One potential explanation is that impulsive individuals experience more intense and frequent positive emotions and therefore report a greater use of suppression of context-inappropriate displays of positive emotions. However, the item that measures impulsivity on the DERS-18, refers specifically to impulsivity *when the respondent is feeling upset*. Therefore, these items better capture negative urgency (i.e., the tendency to act rashly in response to strong negative emotions), rather than positive urgency (i.e., the tendency to act rashly in response to strong positive emotions) or impulsivity in general [36,37]. Given the high correlation between negative and positive urgency [38,39], hypothesizing that individuals chose to suppress given the context, would go against previous research that shows that people high in positive and negative urgency act in the spur of the moment when experiencing intense emotions [37–39]. Our paper suggests differences between positive and negative suppression. Thus, future research could focus on identifying whether these subtypes of suppression are explained by different underlying mechanisms, that could be differentially related to impulsivity based on the intensity of different emotions.

On the ERQ-PN, negative suppression was associated with negative mental health, but to a lesser degree, in comparison to positive suppression. Notably, negative suppression presented a positive correlation with emotional stability, diverging from the negative correlations observed with both positive suppression and overall suppression. This discrepancy may suggest that the context in which suppression is used may influence its outcomes and support distinguishing between positive and negative regulation goals. For instance, in specific contexts, suppressing negative emotions might reflect increased emotional stability, especially in contexts where emotional restraint is valued, as it allows an individual to match the contextual demands [32,40]. However, not all negative suppression is adaptive. For example, when faced with a conflict, expressing anger paired with problem-solving has been shown to be more beneficial than not expressing anger at all [41]. Future research should explore the intersection of multiple emotion regulation strategies to better characterize the process of regulation between reappraisal and suppression. Altogether, these results indicate that the balance between suppression and expression of emotions should be studied in a way that disaggregates between positive and negative emotions to understand how suppression of negative emotions can be helpful in certain contexts.

### 4.3. Clinical Implications

The distinction between positive and negative emotion regulation may carry implications for clinical practice. For instance, interventions focusing on positive affect in depression have been found to be more efficacious than interventions focused on reducing negative affect [42], although this pattern of results is not found consistently across studies [43]. Additionally, integrating a focus on both positive and negative emotion regulation might offer additional benefits in the context of psychological interventions. Geschwind et al. example, found faster symptom improvement when an initial negative-focus cognitive behavioral therapy (CBT) treatment phase was followed by a positive-focus CBT phase.

Future studies could delve into the distinct associations between emotion regulation targets and therapeutic strategies to further our understanding of the nuanced dynamics of emotion regulation. Specific theoretical models, like those proposed by Litz et al. [45], suggest that the emotional numbness and anhedonia often reported by individuals with post-traumatic stress disorder might originate from challenges in expressing positive emotions rather than an inability to experience them. Thus, being able to distinguish positive from negative suppression could be crucial for understanding the regulation of positive emotions in a broader range of mental health disorders, a topic that has received less attention compared to the regulation of negative emotions.

### 4.5. Limitations and Strengths

While the current study provides valuable insights into the role of valence in emotion regulation strategies and mental health outcomes, it is not without limitations. One limitation is the use of a Western sample, as there is evidence that suppression may not have an adverse effect on mental health in an Eastern Asian sample [46]. Thus, differentiating positive from negative suppression in participants from Eastern Asian countries could lead to different results.

Additionally, we relied entirely on self-report measures with one time point. It is possible that differential associations would emerge in an experimental context or with longitudinal data (e.g., individuals may identify moments in which they reappraise to feel positive vs. to feel less negative). Additionally, previous recent research suggests that reappraisal can be done in different ways (e.g., reappraise the emotion vs perspective taking) [47] and for different purposes (e.g., reconstrual vs repurposing) [48] which the ERQ was not designed to capture.

One notable strength of our study lies in the utilization of a large nationally representative sample of online workers, which increases the generalizability of our findings relative to other convenience samples like college students. Another strength is that we use multiple measures of emotion regulation, such as the DERS, alongside the ERQ to establish convergent validity. Also, we included multiple assessments encompassing a wide range of validators including positive and negative dimensions of mental health. Most importantly, we contributed to the field of emotion regulation by studying different targets of emotion regulation in a well-established measure. Our results support the utility of distinguishing between positive and negative suppression and support a commonality of positive and negative reappraisal.

### 4.6. Future Directions

Future research should continue exploring the role of emotional valence’s role in emotion regulation strategies and mental health outcomes. One immediate avenue of research would be to modify the ERQ-PN to make the targets of reappraisal and suppression more consistent across items. Because there is evidence that reappraisal and suppression can change across treatments [49], future work could examine how interventions influence positive and negative reappraisal and suppression distinctively and how these changes relate to changes in outcomes.

Disaggregating between positive vs. negative emotion regulation targets might be especially important for individuals experiencing difficulty expressing positive emotions (e.g., individuals with anhedonia vs. individuals with fear of positive emotions). Additionally, researchers could triangulate the ERQ-PN with other indicators of emotion regulation like text responses and biomarkers like skin conducting. Moreover, the ERQ-PN’s performance in different populations and cultures and its relationship with other measures of emotion regulation strategies should be investigated to further validate its utility as a comprehensive emotion regulation assessment tool.

## 5. Conclusions

In conclusion, the ERQ-PN appears to be a reliable and potentially valid measure of emotion regulation, including positive and negative emotions. The distinction between positive and negative emotions may be necessary for a more refined understanding of emotion regulation and could have important clinical implications. By considering the valence of emotions in emotion regulation strategies, researchers and clinicians can better understand and address the unique challenges individuals face in regulating their emotions and improving their mental health.

### Author Contributions

Conceptualization: Robinson De Jesús-Romero, José R. Chimelis-Santiago, Lauren A. Rutter, Lorenzo Lorenzo-Luaces

Data curation: Robinson De Jesús-Romero, Lorenzo Lorenzo-Luaces

Formal analysis: Robinson De Jesús-Romero, Lorenzo Lorenzo-Luaces

Funding acquisition: Lorenzo Lorenzo-Luaces

Investigation: Robinson De Jesús-Romero, Lorenzo Lorenzo-Luaces, José R. Chimelis-Santiago

Methodology: Robinson De Jesús-Romero, José R. Chimelis-Santiago, Lauren A. Rutter, Lorenzo Lorenzo-Luaces

Project administration: Robinson De Jesús-Romero

Resources: Lorenzo Lorenzo-Luaces

Software: Robinson De Jesús-Romero, Lorenzo Lorenzo-Luaces, José R. Chimelis-Santiago

Supervision: Lorenzo Lorenzo-Luaces

Validation: Lorenzo Lorenzo-Luaces

Visualization: Robinson De Jesús-Romero, Lorenzo Lorenzo-Luaces, José R. Chimelis-Santiago Writing – original draft: Robinson De Jesús-Romero, Lorenzo Lorenzo-Luaces

Writing – review & editing: Robinson De Jesús-Romero, José R. Chimelis-Santiago, Lauren A. Rutter, Lorenzo Lorenzo-Luaces

## Data Availability

Data can be found on the Open Science Foundation (OSF) website: https://osf.io/ezngp/.

## References

1. Gross JJ. The emerging field of emotion regulation: An integrative review. Review of General Psychology. 1998;2: 271–299. doi:10.1037/1089-2680.2.3.271

2. Gross JJ, John OP. Individual Differences in Two Emotion Regulation Processes: Implications for Affect, Relationships, and Well-Being. J Pers Soc Psychol. 2003;85: 348–362. doi:ckv

3. Southward MW, Heiy JE, Cheavens JS. Emotions as context: Do the naturalistic effects of emotion regulation strategies depend on the regulated emotion? J Soc Clin Psychol. 2019;38: 451–474. doi:10.1521/jscp.2019.38.6.451

4. John O, Gross J. Healthy and Unhealthy Emotion Regulation: Personality Processes, Individual Differences, and Life Span Development. J Pers. 2004;72: 1301–1334.

5. Nezlek JB, Kuppens P. Regulating positive and negative emotions in daily life. J Pers. 2008;76: 561–580. doi:10.1111/j.1467-6494.2008.00496.x

6. Beblo T, Fernando S, Klocke S, Griepenstroh J, Aschenbrenner S, Driessen M. Increased suppression of negative and positive emotions in major depression. J Affect Disord. 2012;141: 474–479. doi:10.1016/j.jad.2012.03.019

7. Ruan Y, Reis HT, Zareba W, Lane RD. Does suppressing negative emotion impair subsequent emotions? Two experience sampling studies. Motiv Emot. 2020;44: 427–435. doi:10.1007/s11031-019-09774-w

8. Campbell-Sills L, Barlow DH, Brown TA, Hofmann SG. Acceptability and suppression of negative emotion in anxiety and mood disorders. Emotion. 2006;6: 587–595. doi:10.1037/1528-3542.6.4.587

9. Stellern J, Xiao K Bin, Grennell E, Sanches M, Gowin JL, Sloan ME. Emotion regulation in substance use disorders: a systematic review and meta-analysis. Addiction. John Wiley and Sons Inc; 2023. pp. 30–47. doi:10.1111/add.16001

10. Geisler FCM, Schröder-Abé M. Is emotion suppression beneficial or harmful? It depends on self-regulatory strength. Motiv Emot. 2015;39: 553–562. doi:10.1007/s11031-014-9467-5

11. Galanakis M, Galanopoulou F, Stalikas A. Do positive emotions help us cope with occupational stress? Eur J Psychol. 2011;7: 221–240. doi:10.5964/ejop.v7i2.127

12. Chervonsky E, Hunt C. Suppression and expression of emotion in social and interpersonal outcomes: A meta-analysis. Emotion. 2017;17: 669–683. doi:10.1037/emo0000270

13. English T, John OP. Understanding the social effects of emotion regulation: The mediating role of authenticity for individual differences in suppression. Emotion. 2013;13: 314–329. doi:10.1037/a0029847

14. Gruber J. Can feeling too good be bad? Positive emotion persistence (PEP) in bipolar disorder. Current Directions in Psychological Science. 2011. pp. 217–221. doi:10.1177/0963721411414632

15. Albert DA, Smilek D. Comparing attentional disengagement between Prolific and MTurk samples. Sci Rep. 2023;13. doi:10.1038/s41598-023-46048-5

16. Prolific. Representative samples. 19 Oct 2023.

17. U.S. Census Bureau. 2020 Census Illuminates Racial and Ethnic Composition of the Country. 2022. Available: https://www.census.gov/library/stories/2021/08/improved-race-ethnicity-measures-reveal-united-states-population-much-more-multiracial.html

18. Gratz KL, Roemer L. Multidimensional Assessment of Emotion Regulation and Dysregulation: Development, Factor Structure, and Initial Validation of the Difficulties in Emotion Regulation Scale 1. J Psychopathol Behav Assess. 2004.

19. Victor SE, Klonsky ED. Validation of a Brief Version of the Difficulties in Emotion Regulation Scale (DERS-18) in Five Samples. J Psychopathol Behav Assess. 2016;38: 582–589. doi:10.1007/s10862-016-9547-9

20. Kessler RC, Barker PR, Colpe LJ, Epstein JF, Gfroerer JC, Hiripi E, et al. Screening for Serious Mental Illness in the General Population. Arch Gen Psychiatry. 2003;60: 184. doi:10.1001/archpsyc.60.2.184

21. Prochaska JJ, Sung HY, Max W, Shi Y, Ong M. Validity study of the K6 scale as a measure of moderate mental distress based on mental health treatment need and utilization. Int J Methods Psychiatr Res. 2012;21: 88–97. doi:10.1002/mpr.1349

22. Topp CW, Østergaard SD, Søndergaard S, Bech P. The WHO-5 well-being index: A systematic review of the literature. Psychother Psychosom. 2015;84: 167–176. doi:10.1159/000376585

23. Watson D, Clark LA, Tellegen A. Development and Validation of Brief Measures of Positive and Negative Affect: The PANAS Scales. J Pers Soc Psychol. 1988.

24. Gosling SD, Rentfrow PJ, Swann WB. A very brief measure of the Big-Five personality domains. J Res Pers. 2003;37: 504–528. doi:10.1016/S0092-6566(03)00046-1

25. R Core Team. R: A language and environment for statistical computing. R Foundation for Statistical Computing. 2020. Available: https://www.R-project.org/.

26. Bentler PM. Comparative fit indexes in structural models. Psychol Bull. 1990;107: 238–246. doi:10.1037/0033-2909.107.2.238

27. Rigdon EE. CFI versus RMSEA: A comparison of two fit indexes for structural equation modeling. Struct Equ Modeling. 1996;3: 369–379. doi:10.1080/10705519609540052

28. Hu LT, Bentler PM. Cutoff criteria for fit indexes in covariance structure analysis: Conventional criteria versus new alternatives. Structural Equation Modeling. 1999;6: 1–55. doi:10.1080/10705519909540118

29. Raftery AE. Bayesian Model Selection in Social Research. Sociol Methodol. 1995;25: 111. doi:10.2307/271063

30. Pavlov G, Shi D, Maydeu-Olivares A. Chi-square Difference Tests for Comparing Nested Models: An Evaluation with Non-normal Data. Structural Equation Modeling. 2020;27: 908–917. doi:10.1080/10705511.2020.1717957

31. Zou GY. Supplemental Material for Toward Using Confidence Intervals to Compare Correlations. Psychol Methods. 2007. doi:10.1037/1082-989x.12.4.399.supp

32. Kalokerinos EK, Greenaway KH, Denson TF. Reappraisal but not suppression downregulates the experience of positive and negative emotion. Emotion. 2015;15: 271–275. doi:10.1037/emo0000025

33. Butler EA, Egloff B, Wilhelm FH, Smith NC, Erickson EA, Gross JJ. The Social Consequences of Expressive Suppression. Emotion. 2003;3: 48–67. doi:10.1037/1528-3542.3.1.48

34. Kalokerinos EK, Greenaway KH, Casey JP. Context shapes social judgments of positive emotion suppression and expression. Emotion. 2017;17: 169–186. doi:10.1037/emo0000222

35. Srivastava S, Tamir M, McGonigal KM, John OP, Gross JJ. The Social Costs of Emotional Suppression: A Prospective Study of the Transition to College. J Pers Soc Psychol. 2009;96: 883–897. doi:10.1037/a0014755

36. Whiteside SP, Lynam DR. The Five Factor Model and impulsivity: using a structural model of personality to understand impulsivity. Pers Individ Dif. 2001;30: 669–689. doi:10.1016/S0191-8869(00)00064-7

37. Cyders MA, Smith GT, Spillane NS, Fischer S, Annus AM, Peterson C. Integration of impulsivity and positive mood to predict risky behavior: Development and validation of a measure of positive urgency. Psychol Assess. 2007;19: 107–118. doi:10.1037/1040-3590.19.1.107

38. Cyders MA, Smith GT. Emotion-Based Dispositions to Rash Action: Positive and Negative Urgency. Psychol Bull. 2008;134: 807–828. doi:10.1037/a0013341

39. Cyders MA, Littlefield AK, Coffey S, Karyadi KA. Examination of a short English version of the UPPS-P Impulsive Behavior Scale. Addictive Behaviors. 2014;39: 1372– 1376. doi:10.1016/j.addbeh.2014.02.013

40. Hu T, Zhang D, Wang J, Mistry R, Ran G, Wang X. Relation between emotion regulation and mental health: A meta-analysis review. Psychological Reports: Measures and Statistics. 2014;114: 341–362. doi:10.2466/03.20.PR0.114k22w4

41. Lilienfeld SO, Lynn SJ, Ruscio J, Beyerstein BL. 50 Great Myths of Popular Psychology: Shattering Widespread Misconceptions about Human Behavior. Wiley; 2009. Available: https://books.google.com/books?id=3Nu2EAAAQBAJ

42. Craske MG, Treanor M, Dour H, Meuret A, Ritz T. Positive Affect Treatment for Depression and Anxiety: A Randomized Clinical Trial for a Core Feature of Anhedonia. J Consult Clin Psychol. 2019;87: 457–471. doi:10.1037/ccp0000396

43. Chaves C, Lopez-Gomez I, Hervas G, Vazquez C. A Comparative Study on the Efficacy of a Positive Psychology Intervention and a Cognitive Behavioral Therapy for Clinical Depression. Cognit Ther Res. 2017;41: 417–433. doi:10.1007/s10608-016-9778-9

44. Geschwind N, Arntz A, Bannink F, Peeters F. Positive cognitive behavior therapy in the treatment of depression: A randomized order within-subject comparison with traditional cognitive behavior therapy. Behaviour Research and Therapy. 2019;116: 119–130. doi:10.1016/j.brat.2019.03.005

45. Litz BT, Litz BT, Gray MJ. Emotional Numbing in Posttraumatic Stress Disorder: Current and Future Research Directions. Australian & New Zealand Journal of Psychiatry. 2002;36: 198–204. doi:10.1046/j.1440-1614.2002.01002.x

46. Soto JA, Perez CR, Kim Y-H, Lee EA, Minnick MR. Is expressive suppression always associated with poorer psychological functioning? A cross-cultural comparison between European Americans and Hong Kong Chinese. Emotion. 2011;11: 1450–1455. doi:10.1037/a0023340

47. Webb TL, Miles E, Sheeran P. Dealing with feeling: A meta-analysis of the effectiveness of strategies derived from the process model of emotion regulation. Psychol Bull. 2012;138: 775–808. doi:10.1037/a0027600

48. Uusberg A, Ford B, Uusberg H, Gross JJ. Reappraising reappraisal: an expanded view. Cogn Emot. 2023. doi:10.1080/02699931.2023.2208340

49. Lorenzo-Luaces L, Howard J, De Jesús-Romero R, Peipert A, Buss JF, Lind C, et al. Acceptability and Outcomes of Transdiagnostic Guided Self-help Bibliotherapy for Internalizing Disorder Symptoms in Adults: A Fully Remote Nationwide Open Trial. Cognit Ther Res. 2022. doi:10.1007/s10608-022-10338-5

